# Novel loci and multi-omics risk models for rheumatoid arthritis through a million-participant genome-wide association meta-analysis

**DOI:** 10.64898/2026.06.21.26356058

**Authors:** Galadriel Lucía Velázquez Silva, Jelisaveta Džigurski, Urmo Võsa, Nele Taba, Aare Märtson, Kaspar Tootsi, Katrin Ulst, Raili Müller, Estonian Biobank research team, Elin Org, Triin Laisk, Reedik Mägi, Kristi Läll, Ene Reimann

**Affiliations:** Estonian Genome Centre, Institute of Genomics, University of Tartu, Tartu, Estonia; Tartu University Hospital, Tartu, Estonia

## Abstract

Rheumatoid arthritis (RA) remains incompletely understood, limiting targeted prevention. In this work, genome-wide association study meta-analyses were performed for RA and seropositive RA, comprising approximately one million participants of European ancestry. Eight and six novel genomic risk loci were defined for RA and seropositive RA, and candidate causal genes were identified, highlighting relevant biological pathways, including established immune pathways and estrogen metabolism. Novel disease-specific polygenic risk scores (PRSs) were constructed, enhancing predictive performance over clinical risk factors (incremental C-statistics of 2.7 and 5.1 for RA and seropositive RA, respectively). In parallel, integrating metabolomic data into high-dimensional models enhanced risk stratification over models based on clinical risk factors and genomics, particularly for seropositive RA, where the hazard ratio of the highest decile increased from 4.869 to 5.697. These findings expand the understanding of genetic factors underlying RA and support the value of including PRSs in risk assessment, while suggesting metabolomic integration may further enhance risk stratification, particularly for seropositive RA.

## Introduction

Rheumatoid arthritis (RA) is a systemic autoimmune disease characterised by chronic inflammation that predominantly impacts the joints, affecting approximately 0.5% of the global population, with a generally higher prevalence in industrialised countries and an increasing trend worldwide^1–4^. Consequently, the increasing burden is reflected in elevated rates of disability, morbidity and premature mortality, which, despite therapeutic advances, remain significant^5^. RA is also associated with an increased risk of cardiovascular disease events as well as an elevated risk of infections, depression and malignancy^5–7^. Clinical RA is often preceded by a pre-clinical stage spanning years, characterised by the presence of autoantibodies, resulting in a substantial number of patients already presenting with irreversible joint damage at the time of diagnosis^8,9^. Once RA develops, there is no cure, and treatment can be challenging and requires lifelong therapies^2,5^. Successful management of RA is based on an early diagnosis and a prompt initiation of disease-modifying treatment^5,10,11^. In addition, a significant proportion of patients have difficult-to-treat disease and continue to experience symptoms despite adequate treatment according to current recommendations^5,8,10,11^. RA represents a global public health challenge, and it has become clear that, in addition to the importance of early treatment, prevention or delay of disease onset is also crucial^3,5,8^. Therefore, identifying individuals at elevated risk of developing RA could help guide preventive strategies and early interventions.

Previous large-scale genome-wide association study (GWAS) meta-analyses have identified over 100 risk loci contributing to RA genetic predisposition, with candidate genes pointing to essential roles of the immune system and joint tissues^12,13^. Several studies have shown a strong association between major histocompatibility complex (MHC) genes and RA, particularly for seropositive RA (defined by the presence of rheumatoid factor and/or anti-citrullinated protein antibodies), with the human leukocyte antigen-shared epitope (HLA-SE) being the strongest genetic risk factor, while several non-MHC genes involve more modest associations^4,14,15^. Seropositive RA represents a more homogeneous RA subgroup with a stronger genetic component, distinct disease course and treatment response compared with seronegative RA, which shows greater heterogeneity^16^. This highlights the potential of genetics for understanding the not yet fully elucidated mechanisms underlying RA and its subtypes^16^, and for detecting novel genetic predictors of disease risk and identifying individuals at elevated risk of developing RA^12^.

Beyond genetic predisposition, environmental and lifestyle factors such as smoking, obesity, and microbiome composition further contribute to RA risk in genetically susceptible individuals^4^. Several studies have demonstrated that the use of multiple blood biomarkers can improve the prediction of common diseases and have suggested the added benefits of genetics and metabolomics in conditions like cardiovascular disease and type 2 diabetes^17–20^. Although a prospective study demonstrated improved RA risk stratification by integrating metabolomic profiles and genetic predisposition, the genetic component was based on a limited number of variants from earlier GWAS data, and the differences that might exist among different RA serological subtypes were not examined^21^. Therefore, integrating metabolomics data into risk models may improve early detection and risk stratification for RA and their subtypes, compared with clinical and genetic risk factors alone.

We present a large European GWAS meta-analysis of RA and seropositive RA, comprising approximately one million participants, that identified new genetic risk loci associated with RA and seropositive RA. We characterised novel association signals using a multi-faceted gene prioritisation approach to propose candidate causal genes for both phenotypes. We developed new polygenic risk scores (PRSs) for both phenotypes and evaluated the predictive performance of the PRSs in combination with clinical risk data in the Estonian Biobank (EstBB). Additionally, we developed novel high-dimensional models based on clinical risk factors, genomics and metabolomics data to investigate whether metabolomics can enhance disease risk prediction and stratification over clinical risk factors and genomics-based models.

## Methods

The activities of the EstBB are regulated by the Human Genes Research Act, which was adopted in 2000 specifically for the operations of the EstBB. Individual-level data analysis in the EstBB was carried out under ethical approval 1.1-12/4564 (21.12.2023) from the Estonian Committee on Bioethics and Human Research (Estonian Ministry of Social Affairs), using data according to release application No. 6-7/GI/13482 from the EstBB.

An overview of the study design is shown in Figure 1.

**Figure 1.**
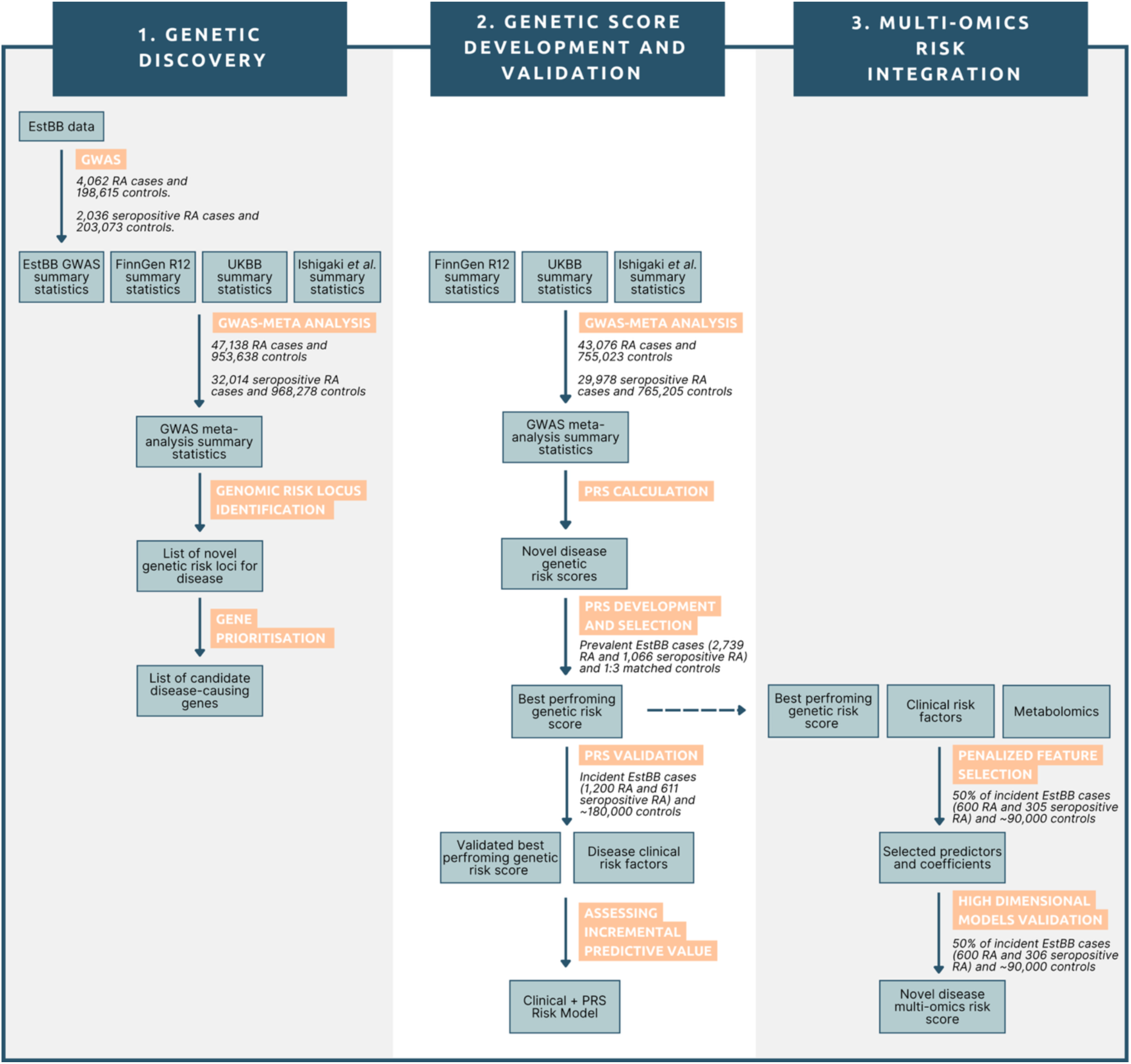
Flowchart of the study design. The study workflow is divided into three main blocks with different objectives: 1) genetic discovery (**left**) with the main aim of identifying novel genetic risk loci associated with rheumatoid arthritis (RA) and seropositive RA, and candidate disease-causing genes; 2) polygenic risk score (PRS) development and validation (**middle**) with the objective of creating and validating disease-specific PRSs and evaluating their incremental predictive value over clinical risk factors; 3) multi-omics risk integration (**right**) with the main goal of integrating genetic, metabolomic, and clinical predictors to develop multi-omics risk models. EstBB, Estonian Biobank; GWAS, genome-wide association study; UKBB, UK Biobank.

### Study cohorts, phenotype definition, genotyping, imputation & quality control

We used data from three European cohorts, the EstBB, FinnGen and UK Biobank (UKBB), and combined these with a previously published meta-analysis performed by Ishigaki et al.^12^ that included 25 additional cohorts with European ancestry (Supplementary Tables 1 and 2).

The EstBB is a volunteer population-based biobank cohort that represents a large portion of the Estonian adult population (∼20 %), with more than 212,000 participants from a wide age range and primarily of European ancestry^22,23^. The EstBB comprises genomics, biomarkers, and health records linked from the databases and registries of national and hospital systems, and additional data about lifestyle and socio-demographic factors, as well as other health-related information collected from self-reported questionnaires^22,23^. All biobank participants (BPs) have signed a broad consent form.

FinnGen is a collaborative project that integrates data from Finnish biobanks and electronic health records. GWAS summary statistics were downloaded from the FinnGen R12 data release (https://r12.finngen.fi) following the instructions outlined here: https://www.finngen.fi/en/access_results. The R12 data freeze included 500,348 individuals (282,064 females and 218,284 males). As all genetic data sets used in this study were based on the Genome Reference Consortium human build 37 (GRCh37), variant positions in the FinnGen summary statistics were lifted from GRCh38 to GRCh37 using a custom script and the database of Single Nucleotide Polymorphism (dbSNP), version 146.

For UKBB, the RA summary statistics were downloaded from the PheWeb browser (https://pheweb.org). The seropositive RA phenotype was downloaded from the GWAS Catalog (study GCST90129457)^24^, which was based on the UKBB field First Occurrences dataset (a dataset that combines diagnoses from multiple sources such as primary care, hospital records, death registers and self-reports).

GWAS summary statistics from the Ishigaki *et al.* dataset^12^ were downloaded from the GWAS Catalog (study GCST90132226) database. This study comprises 22,350 cases and 74,823 controls for RA, and 17,221 cases and 74,823 controls for seropositive RA (male/female ratio was not provided)^12^.

Detailed descriptions of the study cohorts, phenotype definitions, genotyping, imputation, and quality control procedures are provided in the Supplementary Methods and summarised in Supplementary Tables 1 and 2.

### GWAS meta-analysis

Variants with a minor allele frequency (MAF) less than or equal to 0.01, a poor imputation quality (INFO score < 0.8) and/or triallelic variants were removed from all GWAS summary statistics of all different datasets (if data available). Following filtering, a total of 12,927,999 and 10,905,308 variants were retained for RA and seropositive RA, respectively. Inverse variance weighted fixed-effects meta-analyses with genomic control correction using GWAMA software (v2.2.2) were conducted for RA and seropositive RA phenotypes, separately^25^. The RA meta-analysis included 1,000,776 participants of European ancestry (47,138 cases and 953,638 controls), and the seropositive RA meta-analysis included 1,000,292 participants of European ancestry (32,014 cases and 968,278 controls). Associated quantile-quantile plots were generated for both analyses, and the genomic inflation factors (λ) were calculated using a custom script. Variants present in at least 2 out of 4 datasets (EstBB, FinnGen, UKBB and the Ishigaki *et al.* summary statistics) included in the GWAMA meta-analysis were retained for downstream analysis. Variants with a genome-wide significant P-value lower than 5 x 10^-8^ and independent from each other (r² < 0.6) were defined as independent significant SNPs, and those additional independent from each other (at r² < 0.1) were defined as lead SNPs by using Functional Mapping and Annotation of Genome-Wide Association Studies (FUMA) (version 1.7.0)^26^. In addition, genomic risk loci were determined from the grouping of all independent signals (in the same linkage disequilibrium (LD) block based on pre-calculated LD structure derived from the 1000 Genomes Project (1000G) of the European population reference panel) and defining those that are close to each other (< 250 kb) using FUMA. Novel genetic risk loci were defined as those that were not located in a 1 Mb window from any previously associated SNPs with the relevant phenotype according to the GWAS Catalog (accessed 30 October 2025) and using the EFO_0000685 trait ID for RA and EFO_0009459 for seropositive RA. Total SNP-based heritability (*h*²SNP) was estimated using LD score (LDSC) regression^27^ and the HapMap3 reference panel, implemented through the CTG-VL web platform (https://vl.genoma.io), using all imputed autosomal variants with a frequency of the effect allele (EAF) between 0 and 1, including the MHC locus. Observed heritability estimates were transformed to the liability scale using a web-based tool^28^ appropriate for analysing low-prevalence biobank phenotypes (https://medical-genomics-group.shinyapps.io/h2liab/) and using a population prevalence of 0.5% ^1^.

### Gene prioritisation

Gene prioritisation was undertaken for novel loci for RA and seropositive RA. Loci associated with both phenotypes were considered only once in this analysis. Likely causal genes in each novel risk locus were assigned following a prioritisation strategy based on a multifaceted approach (Fig. 2). First, we used the Variant to Gene score (V2G) from Open Targets database (available at https://ftp.ebi.ac.uk/pub/databases/opentargets/genetics/latest/v2g_scored/). V2G score is a single aggregated score that combines multiple data layers, such as quantitative trait loci data (eQTL and pQTL), chromatin interaction and conformation datasets, *in silico* functional prediction (Variant Effect Predictor (VEP)) and distance from the transcript start site (TSS)^29^. The three genes with the highest V2G scores for each novel risk locus were selected. Then, we prioritised genes which have additional evidence in some of the following levels: (1) missense variant in the relevant selected genes, (2) genes with relevant effects (overlapping with disease-related pathways) in mouse models according to the Mouse Genome Database^30^, (3) genes that have the highest Polygenic Priority Score (PoPS) in the genomic risk loci. PoPS assumes that causal genes located near significantly associated SNPs share biological annotations^31^. PoPS scores were calculated in an intermediate step of the fine-mapped locus assessment model of effector genes (FLAMES) pipeline^32^. Disease-related pathways were defined as those pathways that are part of the main affected systems in RA patients, such as the innate immune system, adaptive immune system, cytokine and inflammatory signalling networks, and structural systems (synovial, cartilage, bone).

**Figure 2.**
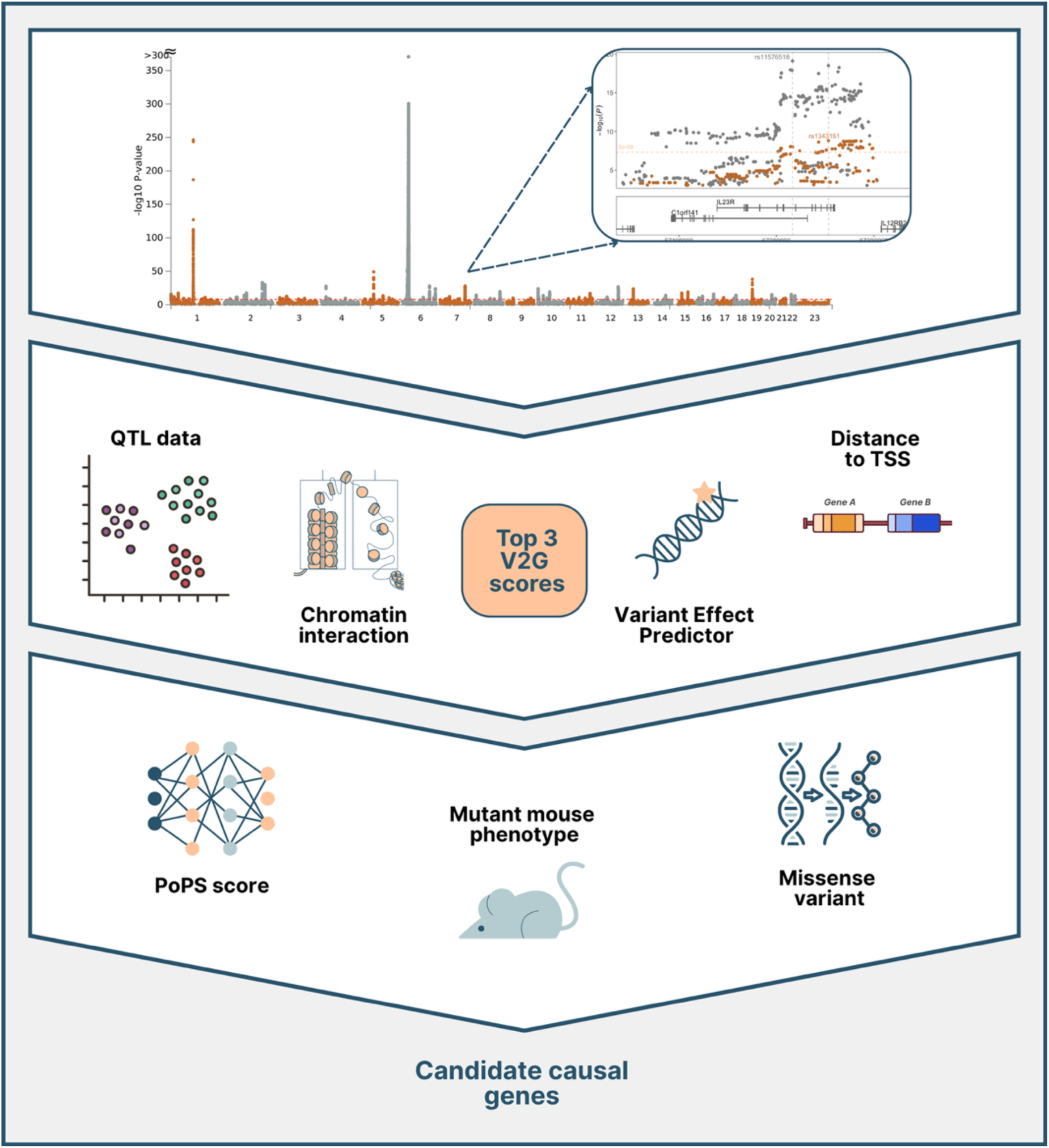
Gene prioritisation strategy for identifying candidate disease-causing genes at novel disease-associated genomic risk loci. Candidate genes were prioritised by integrating several complementary lines of evidence. First, the top three genes with the highest Variant-to-Gene (V2G) score in each novel genomic risk locus were selected. V2G integrates several annotations, including quantitative trait loci (QTL) data, chromatin interaction data, Variant Effect Predictor (VEP), and the distance to the transcription start site (TSS). Second, genes supported by more than one line of evidence were prioritised. Third, genes with only one additional level of evidence were hierarchically weighted and prioritised as follows: (1) disease-related phenotypes in mutant mice; (2) genes with the highest Polygenic Priority Score (PoPS); (3) presence of missense variants or variants in high linkage disequilibrium (r² > 0.6) with coding variants. The images illustrate the strategy and are not representative of real data. Created by the authors using Canva (https://www.canva.com).

Genes with the most additional levels of evidence that were described before and besides the V2G score were first prioritised as the most likely causal genes in each locus. Then, candidate causal genes with only one additional level of evidence were prioritised according to the weight of the evidence level^33^, as follows: (1) relevant phenotype in mouse models; (2) highest PoPS in the genomic risk locus; (3) coding variants.

### PRSs calculation

Leave-one-cohort-out GWAS meta-analyses were performed for RA and seropositive RA by repeating the previous GWAS meta-analyses and leaving out the EstBB cohort (Supplementary Fig. 1). The leave-one-cohort-out GWAS meta-analyses included 43,076 cases and 755,023 controls for RA and 29,978 cases and 765,205 controls for seropositive RA. Genetic variants present in only one dataset, with a MAF < 0.01, low imputation score (INFO score < 0.8), insertions or deletions and/or variants with missing alleles information were excluded. GWAS summary statistics passing quality control were used as development samples for PRS calculation. EstBB genotype and phenotype data were used as an independent target dataset for PRS performance evaluation.

Variant weights were computed using the PRS-CS method (version 1.1.0), applying SNP effect sizes of the development samples for RA and seropositive RA. PRS-CS method ^34^ uses a high-dimensional Bayesian regression framework by placing a continuous shrinkage (CS) which depends on the strength of its GWAS association signal, and preceding on SNP effect sizes for improving the accuracy of the model across genetic architectures. Calculated weights per variant were used for computing individual scores for all BPs using Estonian genotype data (previously filtered by Hardy-Weinberg equilibrium (HWE) of 1 x 10 ^-4^, MAF of 0.001 and INFO score of 0.8). Individual scores were calculated by summing all weighted contributions across the genome for each BP.

### MetaPRS calculation

Phenotype data from the EstBB target dataset were split according to disease status, considering prevalent or incident status depending on whether BPs were diagnosed before or after joining the biobank, respectively. A dataset of prevalent cases and matched controls for each phenotype was defined. Up to 3 controls per case were assigned. Case-control matching was performed by sex and year of birth (within a 5-year range) using the *ccoptimalmatch* package (v0.1.0)^35^ in R (v4.3). The RA dataset included 2,739 cases and 8,215 controls, and the seropositive RA dataset included 1,066 cases and 3,198 controls. We randomly split the prevalent case control dataset into three independent datasets of 40%/20%/40%.

Previously published PRSs (n=3) for RA and for seropositive RA (n=2) were selected from the PGS Catalog^12,36–38^, considering the sample size, ancestry and number of variants included in the development sample used for computing each PRS, as well as the sample size used for evaluating the PRS performance and its performance metrics. The newly developed PRSs and the selected published ones were applied to EstBB genotyped data using the PGS Catalog calculator (pgsc_calc)^39^. Logistic regression models were fitted for each PRS in the first subset (40% of the prevalent cases and controls), adjusting for sex, age and first 10 principal components (PCs). The regression coefficients were extracted from each logistic regression model and pairwise Pearson correlations between the PRSs were calculated in the same dataset.

We derived several combinations of the previously computed PRS and the selected published PRSs into meta scores (metaPRSs), using a regression-based model that calculates the weighted average of the standardised published scores^40^. We excluded combinations simultaneously containing previously computed PRSs, given the high Pearson correlation (0.98) between them. The following formula was used to derive the metaPRSs:

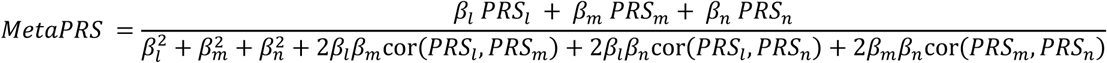

where

- *β_l,m,n_*_…_ are the regression coefficients for each PRS estimated in an independent dataset
- *PRS_l,m,n_*_…_ are the standardised PRSs
- *cor*(*PRS_l_*,*PRS_m_*) is the Pearson correlation between two PRSs in an independent dataset.

All developed metaPRSs were benchmarked using logistic regression adjusted by sex, age and the first 10 PCs in the second independent dataset comprising 20% of prevalent cases and matched controls. Discriminative ability of the metaPRSs was assessed by computing the area under the receiver operating characteristic curve (AUC) in each phenotype and the metaPRS with the highest AUC was selected for downstream analyses.

### Evaluation and comparison of the association of the PRSs with prevalent disease status

All newly developed PRSs and the previously selected metaPRS were additionally benchmarked against the PRSs that were selected from the PGS Catalog for each phenotype by using logistic regression adjusted by sex, age and the first 10 PCs in the third independent dataset (the remaining 40% of prevalent cases and matched controls). As before, the PRS/metaPRS that achieved the highest AUC across case-controls in each phenotype was selected for further analysis. We additionally evaluated the explained variance computing the McFadden pseudo-R^2^, overall odds ratio (OR) and the OR between individuals in each decile of the best-performing PRS, with 95% confidence intervals (CIs), using the same 40% of prevalent cases and matched controls for each phenotype.

### Validation of the PRS in incident disease status

Validation datasets comprised incident cases and unmatched controls (controls that were not previously included in the prevalent case-control datasets), including 1,200 cases and 175,177 controls for RA and 611 cases and 180,316 controls for seropositive RA. The previously selected PRS for each phenotype was further assessed for its predictive ability through computing Harrell’s C-statistic for incident RA and seropositive RA, using Cox proportional hazards models with age as the time scale. Harrell’s C-statistic estimates the probability of concordance between observed and predicted response, ranging from 0.5 (representing performance equivalent to chance) to 1.0 (indicating perfect prediction)^41^. We randomly split the data into 80%/20% for training and testing purposes, respectively.

### Predictive ability of disease-specific PRSs and clinical risk factors in incident disease status

We individually assessed the predictive ability of five clinical risk factors (sex, year of birth, BMI, recent arthralgia and smoking status) and in combination with each other or in combination with the disease-specific PRS. Smoking status was defined as “current”, “former” or “never”. Sex, year of birth, BMI and smoking status were extracted from questionnaires. Recent arthralgia was defined as BPs who experienced joint pain within three years before the baseline (ICD-10 code M25.5), but not due to a prior diagnosis of synovitis and tenosynovitis and/or RA (ICD-10 code exclusions: M65, M05.8, M05.9, M06.0, M06.1, M06.8, and M06.9). The predictive ability of obesity (BMI>30 kg/m²), ever smoked (current or former smokers), and current smokers were additionally individually evaluated.

### Evaluation of the model calibration

We assessed the model calibration over a 5-year follow-up period in the remaining 20% of the incident data (not used for model fitting) for each phenotype. Individual 5-year absolute risks were predicted from the best-performing Cox proportional hazards models combining the selected PRS and clinical risk factors (described above). Standardised incidence ratios (SIRs) were computed using *popEpi* package (v0.4.13)^42^ in R. Expected counts were obtained by applying internally derived reference incidence rates (estimated from the training set of the incident dataset and stratified by age, sex, and calendar year) to the person-years of follow-up in the test set. We additionally reported the SIR by time of follow-up, with 95% CIs. We stratified SIRs by deciles of the predicted risk to examine calibration across decile risk distribution.

### Metabolomic biomarker profiling

Metabolite profiles were generated for all individual blood samples in the EstBB using nuclear magnetic resonance (NMR) spectroscopy, in collaboration with Nightingale Health^23^. Metabolite profiles cover 249 biomarkers (hereafter referred to as extended panel), including low-molecular-weight compounds, lipids and lipoproteins^23^.

### Creating penalised high-dimensional models

NMR metabolomics data were pre-processed by applying missing values to measures that exceeded an outlier threshold of 4 standard deviations from the mean^17^. We randomly split the data (incident cases and controls) into 50%/50% for training and testing purposes. NMR data were log1p-transformed and Z-normalised. Z-normalisation of the metabolomic measures was initially performed in the training dataset. The mean and standard deviation obtained in the training dataset were used to scale the testing dataset. All remaining continuous variables were similarly Z-normalised.

An additional subset of the metabolomic measures was created by selecting 36 CE-marked biomarkers (hereafter referred to as 36 CE-marked panel), that were clinically validated for diagnostic use in Europe^17^.

For RA and seropositive RA, we trained ten models, which all contained

1. baseline age and sex, and additionally included
2. clinical risk factors (BMI, recent arthralgia and current smoking status),
3. top-performing PRS,
4. clinical risk factors, and top-performing PRS,
5. metabolomic measures (36 CE-marked),
6. metabolomic measures (36 CE-marked), and clinical risk factors
7. metabolomic measures (36 CE-marked), clinical risk factors, and top-performing PRS
8. metabolomic measures (extended),
9. metabolomic measures (extended) and clinical risk factors,
10. metabolomic measures (extended), clinical risk factors, and top-performing PRS.

We additionally trained ten models, including interaction terms between the baseline age and sex, the top-performing PRS and all clinical risk factors, to explore whether non-additive effects between predictors could further improve model performance. For models including the 36 CE-marked biomarkers, interaction terms between metabolomic measures and baseline age, sex, the top-performing PRS and all clinical risk factors were also included. Prior to model fitting, we excluded samples that lacked any of the variables needed for score training or prediction. The sample size included in each model is summarised in Supplementary Tables 3 and 4. Models were fitted using Cox proportional hazards regression models with Lasso penalisation (*glmnet* package, version 4.1.7^43,44^ in R), using ten-fold cross-validation and follow-up time as the time scale.

Coefficients were extracted and used for computing single predictor values as the weighted sum of the selected variables and were evaluated in the testing dataset using Cox proportional hazards models and follow-up time as the time scale. Predictive ability of the calculated new scores was quantified by computing Harrell’s C-statistics. We additionally evaluated the hazard ratios (HR) with 95% CIs between individuals in the top decile and the remaining 90% of the study population.

## Results

### GWAS meta-analysis results

We performed a GWAS meta-analysis across four European datasets (Supplementary Tables 1 and 2), including 1,000,776 participants for RA and 1,000,292 participants for seropositive RA. A total of 101 genomic risk loci were identified in RA (399 lead SNPs at genome-wide significance (P <5 x 10^-8^)), and 102 for seropositive RA (416 lead SNPs) (Fig. 3, Supplementary Tables 5 and 6). Of all lead SNPs, a total of 391 and 410 overlapped with previously reported results, leaving a total of eight and six as potential novel findings for RA and seropositive RA, respectively (Fig. 4, Supplementary Tables 7 and 8). Among the eight and six novel genetic risk loci identified for RA and seropositive RA, three loci (rs1267059, rs745328, and rs4281055) were specific to RA, while one locus (rs9782955) was specific to seropositive RA and the remaining loci were shared between both phenotypes. All lead variants of the novel loci were common (MAF > 0.05) and were present in at least 3 out of the 4 datasets included (Fig. 4, Supplementary Tables 7 and 8). The λ values of 1.046 for RA and 1.062 for seropositive RA, along with LDSC intercepts of 1.0094 (95% CI 0.9824 to 1.0364) and 1.0099 (95% CI 0.9827 to 1.0371), respectively, indicate polygenic architecture and no evidence of confounding by population stratification (Supplementary Fig. 2, Supplementary Tables 9 and 10).

**Figure 3.**
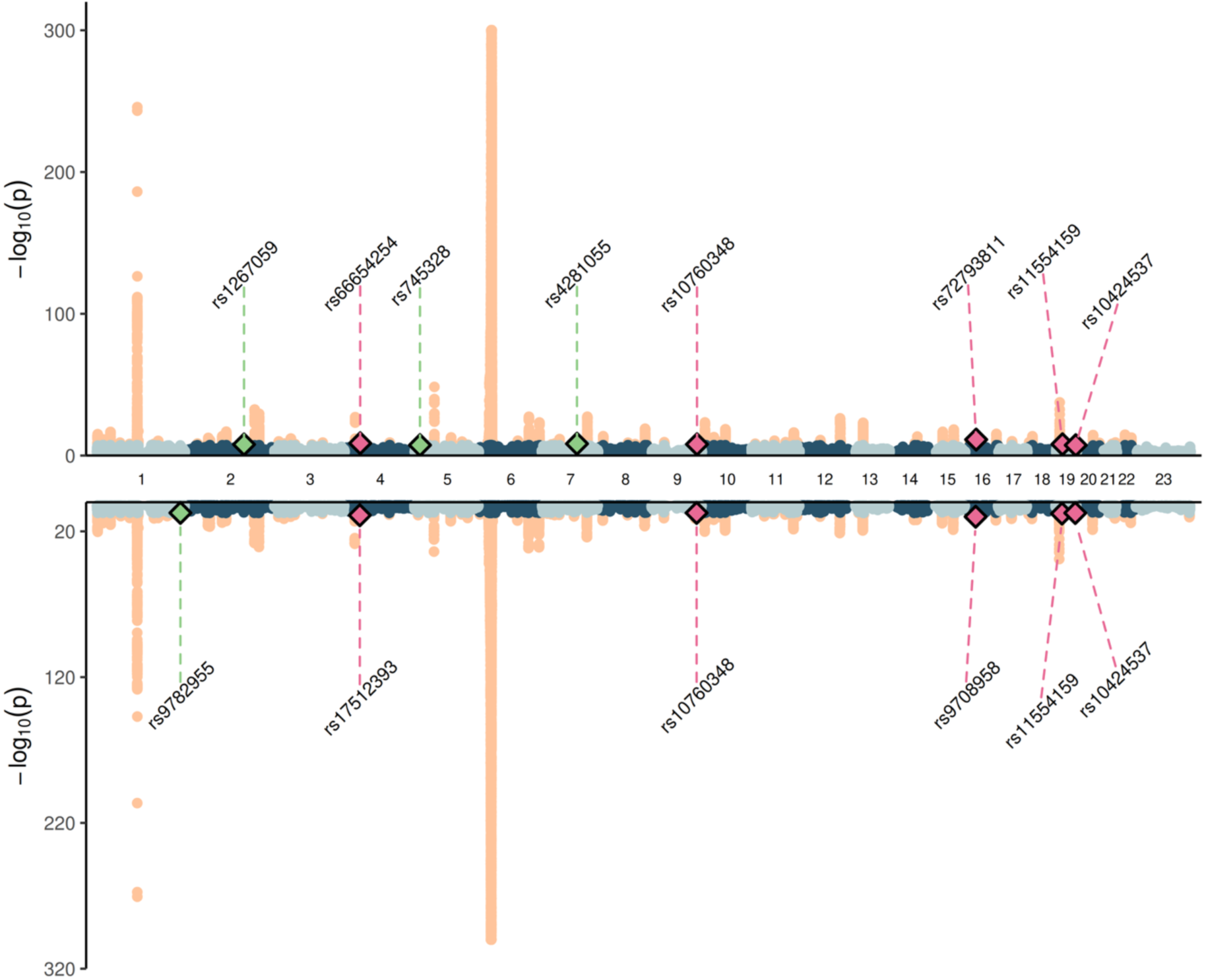
Genome-wide association study meta-analysis results for rheumatoid arthritis phenotypes. Miami plot of the rheumatoid arthritis (RA) (**top**) and seropositive RA (**bottom**) genome-wide association study meta-analyses (*N* = 1,000,776 for RA and *N* = 1,000,292 for seropositive RA). Significant signals (over a Bonferroni corrected p-value of 5 x 10^-8^) are represented in orange, while non-significant signals are represented in blue and grey. Novel loci for both phenotypes are represented in pink, and novel phenotype-specific loci are represented in green.

**Figure 4.**
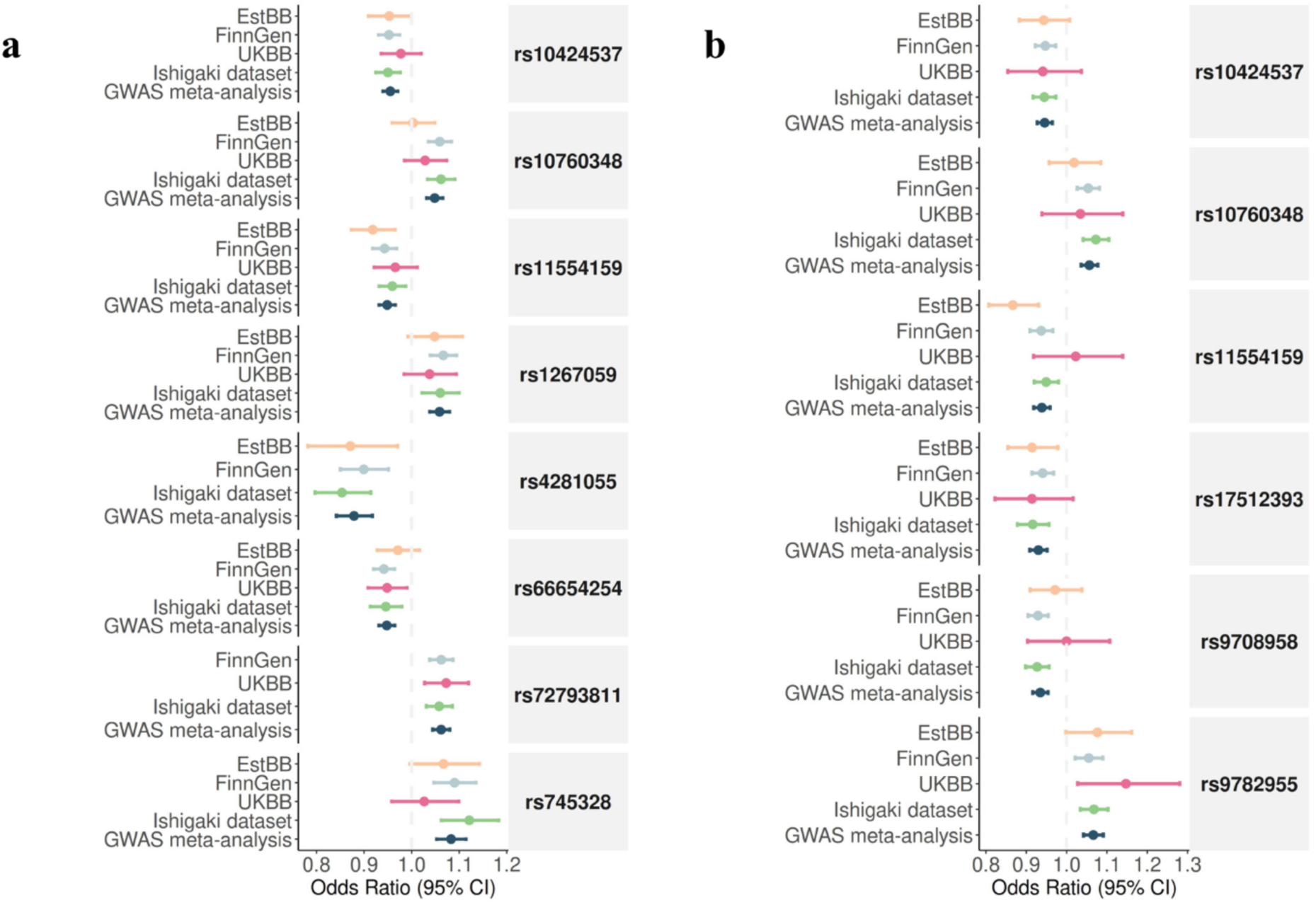
Effect sizes of novel genomic risk loci for rheumatoid arthritis phenotypes from the individual genome-wide association study summary statistics and meta-analysis. Forest plot of the effect sizes (odds ratio), with 95% confidence intervals (CIs) of the novel genomic risk loci associated with rheumatoid arthritis (RA) (**a**) and seropositive RA (**b**) identified in the genome-wide association study meta-analyses (*N* = 1,000,776 for RA and *N* = 1,000,292 for seropositive RA). EstBB, Estonian Biobank; UKBB, UK Biobank; GWAS, genome-wide association study.

### Gene prioritisation

Using a multifaceted approach (Fig. 2) that involves a single score which integrates QTL annotations, chromatin conformation data, *in silico* functional annotations and positional annotations, 27 genes were initially selected as candidate genes for the 9 unique potential novel risk loci associated with one or both phenotypes (Fig. 5). We subsequently identified whether those genes had a relevant phenotype in mouse models, had the highest PoPS score in this disease-associated region or a missense variant in the relevant selected genes, highlighting several putative candidate causal genes for both traits (*RHOH, NEK6, SULT1A1, IFI30, PPP6R1)* as well as RA-specific candidate causal genes (*TANK*, *ANKH*, *FAM105B*, *ARPC1B*) and a seropositive RA-specific candidate causal gene (*LYST*) (Fig. 5). Several of these genes demonstrated phenotypes in mutant mouse models overlapping with RA-related pathways, including *LYST,* which showed broad immune dysfunction with altered levels of cells from the immune and adaptive system, altered cytotoxic function and infection susceptibility, as well as *RHOH,* which showed dysregulated T cell dynamics, altered counts of T cells, lymphocytes and leukocytes and a decreased susceptibility to type I hypersensitivity reactions and *TANK*, which had been linked to an increased B cell activity, immunoglobulins and autoantibodies, lymphoid organ hyperplasia, innate immune dysregulation, elevated pro-inflammatory cytokines and multi-organ inflammation (Fig. 5)

**Figure 5.**
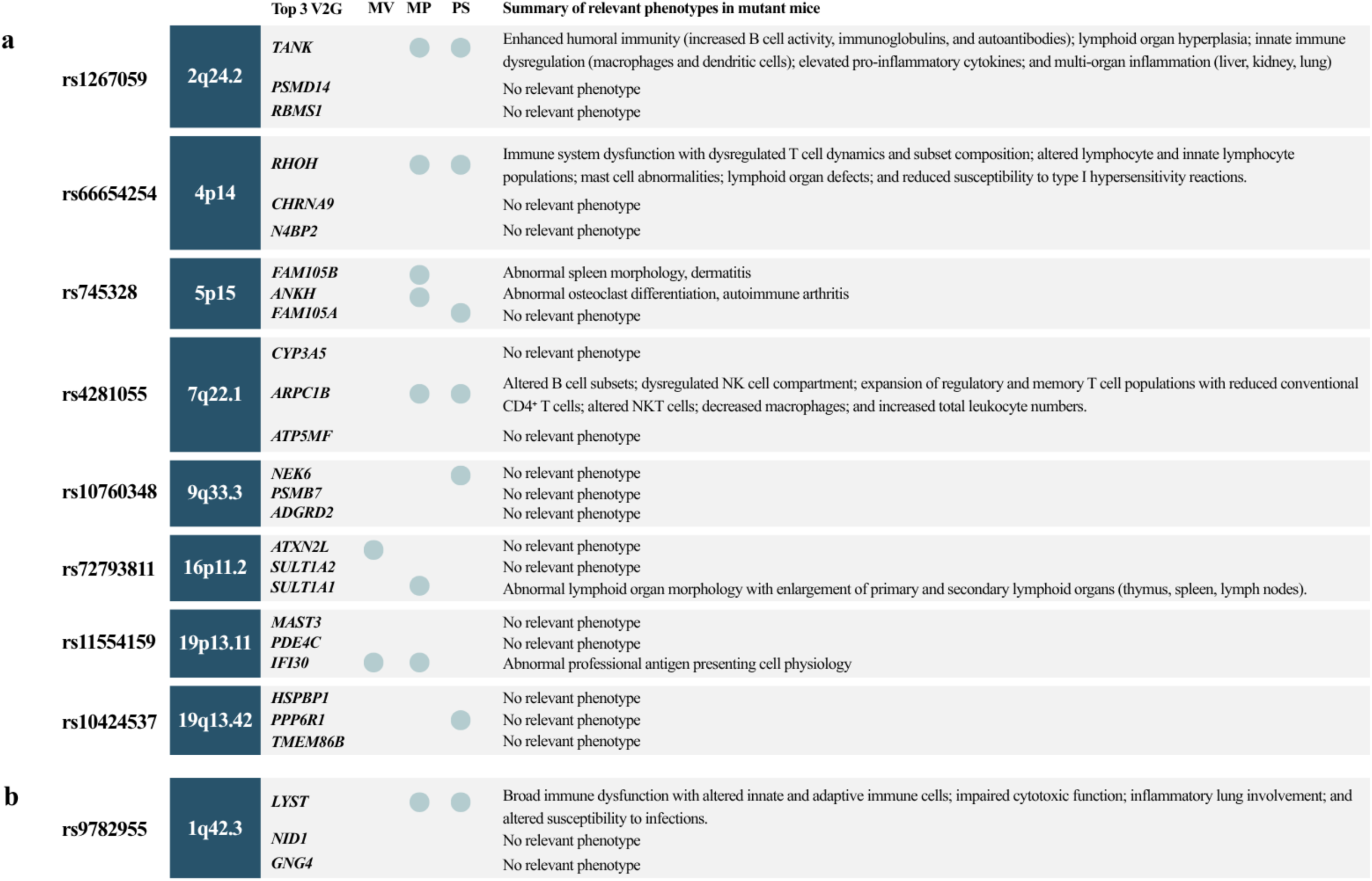
Candidate causal genes. Candidate causal genes for each novel genetic risk locus associated with rheumatoid arthritis (RA) (**a**) and seropositive RA (**b**) based on the gene prioritisation framework. Top 3 V2G, top three genes by Variant-to Gene (V2G) score; MV, missense variant; MP, mouse phenotype; PS, Polygenic Priority Score (PoPS).

### Comparison of different PRSs in association with prevalent disease status

Novel PRSs were computed for RA and seropositive RA using the PRS-CS method and combined with previously published PRSs using a regression-based meta-scoring approach, resulting in a total of 11 novel metaPRSs for RA and 88 for seropositive RA. All metaPRSs were benchmarked by evaluating the case-control discrimination (AUC) in an independent dataset of prevalent cases and matched controls (Supplementary Tables 11 and 12). The metaPRS that achieved the highest AUC was subsequently selected and benchmarked against the individual PRSs in an independent dataset of prevalent cases and controls to identify the overall best-performing PRS (Supplementary Tables 13 and 14). Therefore, the metaPRS created by calculating the optimal weight through combining PGS002088, PGS002745 and the newly developed PRS for RA (Score_RA), achieved the highest discriminative ability among all metaPRSs for both phenotypes separately and was subsequently benchmarked against the selected published PRS in an independent dataset, where it demonstrated the highest discriminative ability, with an AUC of 0.614 (95% CI 0.595 to 0.632) for RA and 0.679 (95% CI 0.650 to 0.708) for seropositive RA. It also showed an OR of 1.31 per 1 standard deviation (SD) (95% CI 1.21 to 1.41) in RA and 1.98 per 1 SD (95% CI 1.81 to 2.16) in seropositive RA. In addition, the McFadden pseudo-R^2^ and OR of each PRS are reported in Supplementary Tables 13 and 14. We also reported the OR with 95% CIs for individuals in each decile of the best-performing score in Supplementary Fig. 3.

### Validation of the best-performing PRS in incident disease

We tested the predictive ability of clinical risk factors and best-performing PRS alone and in combination for incident disease (Fig. 6 and Supplementary Tables 15 and 16). The PRS alone achieved a C-statistic of 0.599 (95% CI 0.579 to 0.619) for RA and 0.669 (95% CI 0.644 to 0.694) for seropositive RA. For RA, the year of birth was the strongest individual predictor among the individual clinical predictors (0.657, 95% CI 0.638 to 0.676), followed by sex (0.604, 95% CI 0.587 to 0.621), smoking status (0.540, 95% CI 0.520 to 0.560), recent arthralgia (0.534, 95% CI 0.514 to 0.554) and BMI (0.525, 95% CI 0.505 to 0.545). For seropositive RA, year of birth showed the highest C-statistic among the individual clinical predictors (0.640, 95% CI 0.613 to 0.667), followed by sex (0.638, 95% CI 0.614 to 0.662), smoking status (0.577, 95% CI 0.549 to 0.605), recent arthralgia (0.569, 95% CI 0.542 to 0.596) and BMI (0.567, 95% CI 0.540 to 0.594). By adding the PRS to all clinical risk factors (Fig. 6 and Supplementary Tables 15 and 16), an increment of 2.7 and 5.1 units was observed for RA and seropositive RA, achieving a C-statistic of 0.731 (95% CI 0.714 to 0.748) and 0.750 (95% CI 0.729 to 0.771) compared with all clinical risk factors, which reached a C-statistic of 0.704 (95% CI 0.686 to 0.722) and 0.699 (95% CI 0.675 to 0.723), respectively.

**Figure 6.**
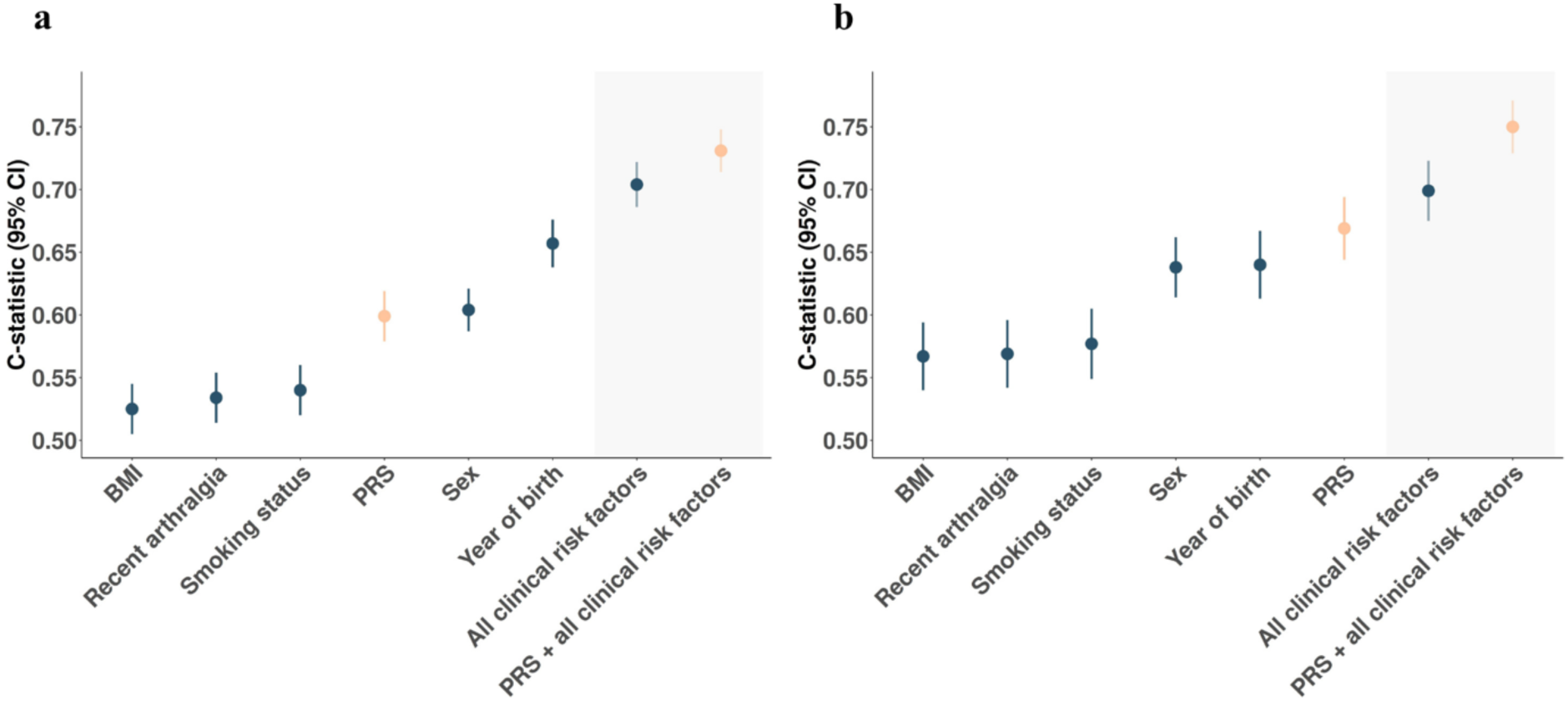
Predictive ability of disease-specific polygenic risk scores and clinical risk factors for rheumatoid arthritis phenotypes. Predictive performance (C-statistic) of disease-specific polygenic risk scores (PRSs) and clinical risk factors, along with the 95% confidence intervals (CIs) of Cox proportional hazards models, adjusted for first 10 principal components for incident rheumatoid arthritis (RA) (**a**) and incident seropositive RA (**b**) in the Estonian Biobank. Clinical risk factors and PRSs are modelled individually on the left side of the plots (white area) and jointly on the right side (grey area).

The best-performing model (PRS + all clinical risk factors) was subsequently evaluated in the independent test set (20% of incident data), achieving a C-statistic of 0.728 (95% CI 0.691 to 0.765) for RA and 0.722 (95% CI 0.669 to 0.774) for seropositive RA.

### Evaluation of model calibration

We evaluated the calibration of the best-performing model (PRS × sex + year of birth + BMI + smoking status + recent arthralgia) for each phenotype, over 5 years of follow-up. We calculated the SIRs using the *popEpi* package in R. Overall SIR standardised by age, sex and calendar year was 1.07 (95% CI 0.9 to 1.26) for RA and 0.93 (95% CI 0.72 to 1.19) for seropositive RA. SIRs by time of follow-up and by predicted risk decile are reported in Supplementary Fig. 4.

### Selecting informative predictors for creating high-dimensional models

Metabolomic biomarkers for all BPs (*N ∼* 200,000) were measured by NMR spectroscopy in blood samples, which were collected at EstBB recruitment time (baseline time). We trained several Cox proportional hazards models with Lasso penalisation and tenfold cross-validation to predict the incidence of RA and seropositive RA in half of the dataset of incident cases and controls from EstBB. We allowed the models to select the most informative variables from the metabolomics data (full version and a subset of 36 CE-marked metabolites), clinical risk factors (sex, age at recruitment, BMI, recent arthralgia and smoking status) and the best-performing PRS for each phenotype. All selected variables and their Lasso coefficients in each model are reported in Supplementary Tables 17 and 18. Baseline age and sex were not fixed covariates but were consistently selected across all RA models, and baseline age was consistently selected in all seropositive RA models. For both phenotypes, PRS was consistently selected across all models in which it was included (Supplementary Tables 17 and 18). Clinical risk factors-based models selected BMI at baseline, smoking status at baseline and recent arthralgia for RA, while fewer variable were selected for seropositive RA (Supplementary Tables 17 and 18). Metabolomics-based models retained a substantially larger number of biomarkers. A core subset was consistently selected across all models, particularly markers of systemic inflammation and general metabolic status, including glycoprotein acetylation (GlycA), albumin, and creatinine, as well as amino acids (histidine and valine) (Supplementary Tables 17 and 18). In addition, a broader set of metabolites was frequently selected for both phenotypes, such as markers involved in fatty acid composition (saturated fatty acids (SFA%), docosahexaenoic acid (DHA%), omega-6-related markers), amino acids (alanine, phenylalanine), glycolysis and energy metabolism (glucose, lactate, pyruvate, β-hydroxybutyrate, acetone) and lipoprotein subclass measures (HDL/VLDL/LDL). For the 36-marker panel, approximately one-third to one-half of biomarkers were selected, whereas models based on the extended metabolomics panel retained a broader set of lipoprotein and metabolic intermediates for both phenotypes.

### Comparison of hazard ratios for incident disease among metabolomic, genetic, clinical risk factor-based and combined scores

Single predictor values were computed as the weighted sum of the previously selected variables for each model, and their performances were evaluated using Cox proportional hazards models in an independent dataset (half of the dataset of incident cases and controls) (Supplementary Tables 19 - 22). As a similar performance was observed in models both with and without interaction terms, results without interaction terms are presented. Fig. 7 illustrates the performance of the scores by comparing the relative risk of the top decile versus the remaining population estimated from Lasso-penalised Cox proportional hazards models for incident RA and seropositive RA in EstBB. The multi-omics scores (based on clinical risk factors, genomics and metabolomics) outperformed the models based individually on clinical risk factors, genomics or metabolomics alone, for both phenotypes (Fig. 7, Supplementary Tables 23 and 24). Notably, multi-omics scores further improved risk stratification (Fig. 7). For incident RA, the HR of the highest decile versus the remaining population increased from 2.882 (95% CI 2.661 to 3.103) in models based on clinical risk factors and genomics to 3.060 (95% CI 2.842 to 3.278) when metabolomics (extended panel) was added. Similarly, for seropositive RA, the HR increased from 4.869 (95% CI 4.599 to 5.139) to 5.697 (95% CI 5.432 to 5.962) (Fig. 7, Supplementary Tables 23 and 24).

**Figure 7.**
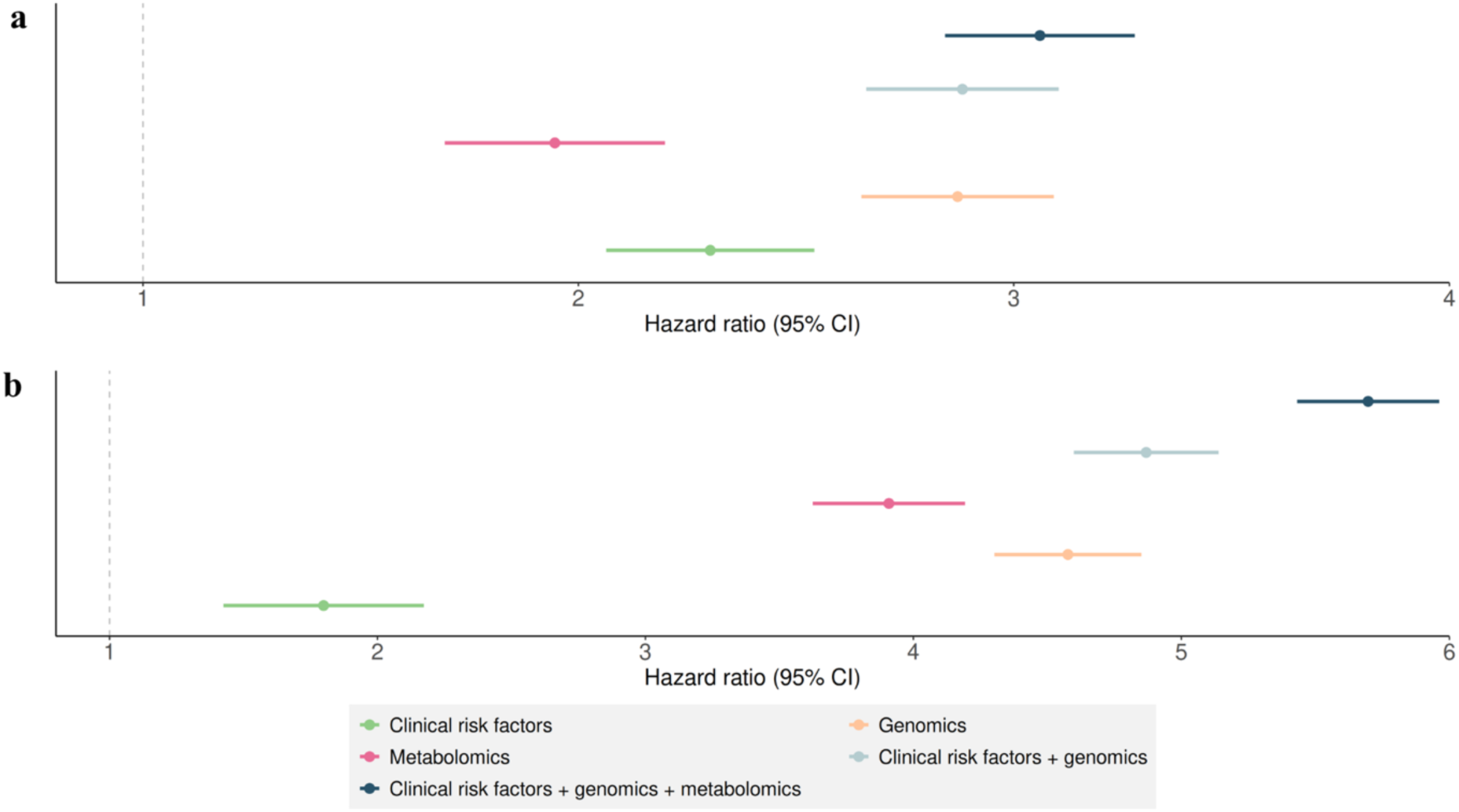
Risk stratification of high-dimensional models for rheumatoid arthritis phenotypes. Hazard ratios (HRs) with the 95% confidence intervals (CIs) of the highest risk decile versus the remaining population, estimated using Lasso-penalised Cox proportional hazards models for incident rheumatoid arthritis (RA) (**a**) and seropositive RA (**b**), in Estonian Biobank. Models included disease-specific polygenic risk scores (genomics), clinical risk factors, biomarkers identified by nuclear magnetic resonance spectroscopy from a panel of 249 biomarkers (metabolomics) and their combinations.

## Discussion

This study represents one of the largest RA and seropositive RA GWAS meta-analyses conducted to date, encompassing over one million participants and including 47,138 RA cases and 32,014 seropositive RA cases. Although previous studies reported a similar total sample size^12,13^, our study includes a substantially higher number of cases (representing roughly double the number of cases of previous studies), resulting in a higher effective sample size. By integrating large-scale population biobanks with previously established cohorts, our study increased statistical power, enabling more efficient discovery.

Among the 101 and 102 genetic risk loci identified in the current study, eight and six were identified as novel findings for RA and seropositive RA, respectively, of which three and one were specific to RA and seropositive RA, respectively. Using a comprehensive mapping approach combining multiple levels of evidence, several putative genes that share disease-relevant pathways were prioritised as potential causal genes for RA and seropositive RA. Genes with the highest V2G score were first selected, integrating functional and regulatory evidence, followed by prioritisation of genes with missense variants, disease-relevant mouse phenotypes or high PoPS scores, highlighting *RHOH, NEK6, SULT1A1, IFI30, PPP6R1* as candidate causal genes of both phenotypes, as well as RA-specific candidates (*TANK*, *ANKH*, *FAM105B*, *ARPC1B*) and a seropositive RA-specific candidate (*LYST*).

Among these loci, our findings support the role of previously reported candidate genes, including *RHOH*, which was identified by Song et al.^45^ and found to be upregulated in RA synovial tissue in the same study, reinforcing its involvement in disease-relevant immune pathways.

Our study also highlighted several candidate causal genes that have not been previously associated with RA in GWAS and converge on NF-κB dysregulation and innate immune pathways, central pathways in RA pathogenesis^46^. NF-κB regulates the expression of pro-inflammatory cytokines, playing a role in driving synovial inflammation in RA^46^. Among the identified candidate genes, *TANK* (TRAF Family Member-Associated NF-κB Activa tor) negatively regulates NF-κB^47^. In addition, inhibition of the *TANK* downstream kinase reduced antibody-dependent arthritis in collagen-induced arthritis mouse models, with minimal effects on antibody-independent inflammation^48^. *PPP6R1*, encoding the regulatory subunit of protein phosphatase 6 (PP6), has a known role in the negative regulation of NF-κB signalling and regulating lymphocyte function and has been shown to be differentially methylated and expressed in RA synovium^49,50^. *FAM105B* (*OTULIN*) is a key negative regulator of NF-κB and inflammatory signaling through its role in removing linear ubiquitin chains, and defects in this protein are associated with inflammation and autoimmunity^51^.

Several novel candidate causal genes converge on pathways central to antigen processing and immune activation. *IFI30* has strong biological support as it encodes the lysosomal thiol reductase GILT protein, which is a key regulator of MHC class II antigen processing, a central pathway for antigen presentation and T cell activation in RA^52^. In addition, proteomic analyses have shown that IFI30 protein levels changed significantly over 15 years prior to disease onset, suggesting a role in early pathogenic processes^53^. *LYST*, encoding a lysosomal trafficking regulator, has been associated with seropositive RA in our study. Functional studies have shown that it regulates endolysosomal trafficking and TLR3/TLR4-mediated TRIF signalling, pathways involved in antigen processing and innate immune activation, relevant to autoimmune responses in RA^54^. Furthermore, a recent DNA methylation study identified *LYST* among blood-based epigenetic markers potentially informative for RA diagnosis^55^. *LYST* may represent potential mechanistic differences between RA subtypes, warranting further investigation.

Other candidate genes are involved in additional biological processes relevant to RA pathogenesis. *SULT1A1* (Sulfotransferase Family 1A Member 1), which encodes a key enzyme responsible for metabolising drugs, hormones and xenobiotics and with a role in estrogen metabolism, may contribute to RA susceptibility, given that fluctuations in estrogen levels across menopause, pregnancy, childbirth and breastfeeding have been suggested to modulate RA risk^56^. *ANKH* encodes a multipass transmembrane protein that regulates extracellular pyrophosphate homeostasis, a key process in tissue calcification, and has additionally been linked to a progressive generalised form of arthritis in mouse models^57^. *ARPC1B* encodes a component of the Arp2/3 complex that regulates actin cytoskeletal dynamics, and studies have shown that loss-of-function mutations in *ARPC1B* cause immunodeficiency with immune dysregulation and autoimmune manifestations^58^, including chronic arthritis with rheumatoid factor positivity^59^. *NEK6* (NIMA-related kinase 6) is a serine/threonine kinase best characterised in cell cycle regulation. Although limited evidence supports its possible relevance to disease, *NEK6* expression has been reported in peripheral blood mononuclear cells from DMARD-naive patients who have not received treatment yet, with similar upregulation also observed in arthritic mice^60^.

These novel findings reinforce the roles of established pathways in the pathogenesis of RA, such as NF-κB signalling, antigen processing and immune activation and cytoskeletal regulation in T cell activation. In addition, this study highlights candidate causal genes involved in biological processes such as estrogen metabolism and pyrophosphate-mediated joint tissue homeostasis, processes that have been shown to be relevant to RA through experimental and epidemiological studies^56,57,61,62^. However, follow-up analyses, including functional validation or Mendelian randomisation analysis, are needed to confirm these candidate causal genes and the potential biological pathways involved in the pathogenesis of RA and seropositive RA.

Our study also presents novel disease-specific PRS that outperformed previous ones in EstBB. The RA PRS created in this study was the third-best individual predictor for incident RA and the best individual predictor for incident seropositive RA when compared with sex, year of birth, BMI, recent arthralgia and smoking status. The addition of disease-specific PRSs to disease-specific clinical-based models improved predictive ability for incident RA (C-statistic of 0.731) and incident seropositive RA (C-statistic of 0.750) compared with the five clinical risk factors alone (C-statistic of 0.704 for RA and 0.699 for seropositive RA).

In high-dimensional risk models, markers of systemic inflammation and general metabolic status, including GlycA and albumin, were consistently selected as informative predictors across all models incorporating metabolomic data, in agreement with their previously reported associations with incident RA risk^21^. Although the integration of the metabolomic data layer modestly enhanced model performance relative to clinical risk factors and genomics, the addition of metabolomics increased the HR of the highest decile compared with the remaining population (3.060 for RA and 5.697 for seropositive RA), suggesting a potential improvement in risk stratification for both phenotypes, over model based on genomics and clinical data (2.882 and 4.869 for RA and seropositive RA, respectively). These findings are consistent with a previous study^21^ suggesting improved overall RA risk stratification when integrating unfavourable NMR metabolomic profiles and RA genetic susceptibility, while our findings suggest a greater benefit specifically for seropositive RA. Nevertheless, the evaluation of whether the observed improvement could be translated into clinical utility remains to be established.

Several limitations of this study should be acknowledged. First, GWAS meta-analysis and PRS construction were based exclusively on European ancestry populations, potentially limiting the generalisability of the findings to non-European populations. Second, phenotype definitions varied across cohorts possibly introducing heterogeneity and potentially affecting the reliability of the associations. Third, metabolite levels were measured and included in risk prediction models at baseline, not accounting for potential changes over time, which may limit the predictive ability of the metabolomic models.

One of the key strengths of this study is the use of EstBB data, comprising approximately the 20% of the adult population, including NMR data, phenotype questionnaire and follow-up data from national health-related registries, making EstBB data well suited for validating PRS models and creating multi-omics risk prediction models.

In summary, this study represents one of the largest GWAS meta-analyses for RA and seropositive RA to date, identifying novel genetic risk loci for both phenotypes, proposing several candidate causal genes for RA and specifically for seropositive RA, with functions in RA-relevant pathways, including established immune pathways such as NF-κB signalling, antigen processing and immune activation, and cytoskeletal regulation in T cell activation, and processes such as estrogen metabolism and pyrophosphate-mediated joint tissue homeostasis. Additionally, we presented novel disease-specific PRSs and highlighted the potential to improve predictive ability when adding PRS on top of clinical risk factors. Lastly, we constructed multi-omics risk prediction models with the potential to improve RA risk stratification beyond clinical and genomic factors, particularly for seropositive RA.

## Supporting information

Supplementary methods and five supplementary figures

## Data Availability

Pseudonymized data and/or biological samples can be accessed for research and development purposes in accordance with the Estonian Human Genome Research Act. To access data, the research proposal must be approved by the Scientific Advisory Committee of the Estonian Biobank as well as by the Estonian Committee on Bioethics and Human Research. Access to samples requires the same approval process and an additional approval from the Senate of the University of Tartu.

## Acknowledgements

This project has received funding from the Ministry of Education and Research Centres of Excellence grant TK214 Centre of Excellence for Personalised Medicine, and by the Estonian Research Council grants PSG776, PRG1911, PRG1414, LP1GI19572; and the European Union’s Horizon Europe research and innovation programme under grant agreement No 101095084 and No 101060011 and through the European Regional Development Fund (Project No. 2021-2027.1.01.24-0444). Views and opinions expressed are, however, those of the author(s) only and do not necessarily reflect those of the European Union or European Research Executive Agency. Neither the European Union nor the granting authority can be held responsible for them.

The research was conducted using the Estonian Centre of Genomics/Roadmap II funded by the Estonian Research Council (project number TT17).

Data analysis was carried out in part in the High-Performance Computing Centre of the University of Tartu.

We thank the participants in the EstBB, FinnGen, UKBB and the Ishigaki cohorts for their valuable contributions.

This study was mostly written during the writing retreats organised by the Institute of Genomics of the University of Tartu.

## Author contribution

R.Mägi and E.R. designed the study. G.L.V.S. performed the GWAS, GWAS-meta analysis, GWAS follow-up, PRS calculation, PRS follow-up analysis, and multi-omics analysis. N.T. performed metabolomics data preprocessing. J.D. and U.V. contributed to PRS analysis. E.O., T.L., R.Mägi, E.R. and K.L. supervised the analyses. K.T., K.U. and R.Müller provided clinical context and clinical interpretation of the findings. A.M., L.M., T.M., R.Mägi, M.M., M.N. and G.H., from the EstBB Research Team, collected, genotyped, performed the quality control and imputed the EstBB data. P.P., N.T., E.A., J.K. and U.V. from the EstBB Research Team performed the metabolomics layer generation, development and QC of the data. G.L.V.S. wrote the first draft of the manuscript. All authors critically reviewed the paper.

