## Supplementary methods and five supplementary figures for "Novel loci and multi-omics risk models for rheumatoid arthritis through a million-participant genome-wide association meta-analysis"

#### *Phenotype definition*

Phenotype definitions are summarised in Supplementary Tables 1 and 2.

Estonian Biobank (EstBB) rheumatoid arthritis (RA) cases were defined with the following codes from the International Classification of Diseases (ICD-10 codes): M05.8 “Other seropositive rheumatoid arthritis”, M05.9 “Seropositive rheumatoid arthritis, unspecified”, M06.0 “Seronegative rheumatoid arthritis”, M06.1 “Adult-onset Still disease”, M06.8 “Other specified rheumatoid arthritis” and/or M06.9 “Rheumatoid arthritis, unspecified”, obtained from the Estonian National Health Insurance Fund and with at least one diagnosis made by a rheumatologist. Seropositive RA cases were defined using the ICD-10 M05.8 “Other seropositive rheumatoid arthritis” and/or M05.9 “Seropositive rheumatoid arthritis, unspecified” with at least one diagnosis made by a rheumatologist. Controls were defined as biobank participants (BPs) who did not have the respective ICD code assigned for cases. BPs who have the relevant ICD-10 but were not diagnosed by a rheumatologist were excluded.

FinnGen RA cases (R12 release)[1] were defined using the FinnGen endpoint “M13_RHEUMA” (https://r12.finngen.fi/pheno/M13_RHEUMA) based on the ICD-10 code M05 “Seropositive rheumatoid arthritis” or M06 “Other rheumatoid arthritis”, from hospital discharge or cause-of-death registries, and controls were individuals without RA and not included in the arthropathies FinnGen endpoint “M13_ARTHROPATHIES”. Seropositive RA cases were defined using the FinnGen endpoint “RHEUMA_SEROPOS_WIDE”, corresponding to the ICD-10 M05 “Seropositive rheumatoid arthritis”, and controls were case-free individuals with no diagnoses included in the broader seropositive RA endpoint “RHEUMA_SEROPOS” (https://r12.finngen.fi/pheno/RHEUMA_SEROPOS_WIDE), corresponding to the ICD-10 M05 “Seropositive rheumatoid arthritis”.

UK Biobank (UKBB) RA cases were defined using PheCode classification (PheCode 714.1), while seropositive RA cases were defined by Zorina-Lichtenwalter K *et al.*[2] as participants with any evidence of the ICD-10 code M05 "Seropositive rheumatoid arthritis" from any of the sources used for creating the First Occurrence dataset (a dataset which combines diagnoses from multiple sources such as primary care, hospital records, death registers and self-reports)[2].

RA cases in Ishigaki *et al.* were defined either by using the 1987 American College of Rheumatology (ACR) criteria or the 2010 ACR/European League Against Rheumatism criteria or were diagnosed with the relevant disease by a rheumatologist[3]. Seropositive cases were defined as RA cases with seropositive status available[3].

***Genotyping, imputation & quality control***

Genotyping, imputation, and quality control procedures are summarised in Supplementary Tables 1 and 2.

Estonian Biobank participants were genotyped using four revisions of the Illumina GSA array (GSAMD-24v1, GSAMD-24v2, ESTchip-1_GSAv2, ESTchip-2_GSAv3) across 17 genotyping batches, each containing a minimum of approximately 1,000 samples. All quality control procedures were performed for each batch separately. At the sample level, individuals with a call rate <95% and/or a sex mismatch between genotype and phenotype data were excluded. At the variant level, genotypes were removed if they failed one of the following filters: minimum call rate 95%, Hardy–Weinberg equilibrium exact test p-values >1e-4, Illumina cluster separation >0.4, and Illumina GenTrain score >0.6. Genotyping data was processed using the GRCh37 human genome reference. To control for potential genotyping batch effects, variants showing inconsistent allele frequencies across batches were removed. Specifically, variant allele frequencies were calculated for each genotyping batch containing more than 10,000 samples (9 batches in total). The mean allele frequency across these batches was then computed, and variants with frequencies deviating by more than 5% from the mean in any batch were excluded from the combined dataset. Only single-nucleotide variants (SNVs) were retained for imputation. Among these, AT, GC, and rare variants (minor allele frequency <1%) were removed. For SNVs typed by more than one probe, genotype concordance between different probes within each sample was assessed, and variants with discordant calls were removed on a per-sample basis. This was followed by another round of a variant-level 95% call rate filter. After quality control, all genotyping batches were merged into a single cohort dataset. Approximately 307K SNVs passed all quality control filters and were used for phasing and imputation. The dataset was phased reference-free using Eagle v2.4.1. Subsequent imputation of the pre-phased data was performed with Beagle v5.4 (beagle.22Jul22.46e.jar), using a population-specific haplotype reference panel derived from whole-genome sequencing data of 2,056 Estonian individuals. Association analyses were carried out by using REGENIE (v3.2) software[4], implementing a mixed-model-based approach using sex, year of birth and the first 10 principal components (PCs) data as covariates, including 4,062 cases and 198,615 controls for RA and 2,036 cases and 203,073 controls for seropositive RA (Supplementary Fig. 5).

FinnGen R12 samples were genotyped using Illumina and Affymetrix (Thermo Fisher Scientific) arrays and subsequently went through a quality control based on removing individuals with high genotype missingness (>5%), sex mismatch, excess heterozygosity (± 4 standard deviations) and variants with high missingness (>2%), low Hardy-Weinberg disequilibrium ($P < 1\times10^{-6}$) or minor allele count <3[1]. Prior to imputation, genotypes were pre-phased using Eagle v2.3.5 (https://finngen.gitbook.io/documentation/methods). Genotypes were then imputed using the Finnish-specific Sequencing Initiative Suomi (SISu) v.4.2 imputation reference panel (https://finngen.gitbook.io/documentation/methods). This panel is based on high-coverage (25x) whole-genome sequencing data for 8,554 Finnish individuals (https://finngen.gitbook.io/documentation/methods). A post-imputation quality control was conducted by analysing the distribution of imputation INFO values, Minor Allele Frequency (MAF) differences between the target dataset and the reference panel and ensuring chromosomal continuity of imputed genotypes (https://finngen.gitbook.io/documentation/methods). Samples with non-Finnish ancestry as well as duplicated/twins were removed. Association analyses were run by using REGENIE (v.2.2.4)[4] with sex, age, the first 10 PCs and genotyping batch as covariates (https://finngen.gitbook.io/documentation/methods). Genetic relatedness was accounted for by the model.

A custom-made Affymetrix chip was used for genotyping the UKBB samples, UK BiLEVE Axiom in the first 49,950 participants and with Affymetrix UKBB Axiom array in the remaining participants, with 95% of the signals shared between the two arrays[5]. Two datasets derived from the UKBB were included in this study. The first was the UKB-SAIGE dataset, comprising 409,728 individuals of White British ancestry. Variants failing quality control in more than one batch, with a missing rate > 5% or a MAF < 0.0001, as well as samples that were identified as outliers for heterozygosity and missingness, were removed[5]. The Haplotype Reference Consortium (HRC) panel was used to impute all individuals, resulting in more than 20 million variants (https://pheweb.org/UKB-SAIGE/about). A generalized mixed model association test that employs a saddle point approximation to address case-control imbalance called SAIGE was used for binary outcome analyses (https://pheweb.org/UKB-SAIGE/about). The analyses were adjusted for genetic relatedness, sex, birth year and the first four PCs.

The second dataset derived from the UKBB was the First Occurrences dataset and has been previously described[2]. Briefly, imputation was based on the UK10K reference panel[2]. Variant quality control included heterozygosity rate |Fhet| ≤ 0.2, call rate >0.95, Hardy–Weinberg equilibrium P > $1\times10^{-8}$, and minor allele frequency >0.01, while sample quality control excluded mismatches between reported and genotyped sex. The association test was performed using REGENIE[4], adjusting for age, sex, and 10 genetic PCs with genetic relatedness accounted for within the model[2].

Details of the genotyping platforms, imputation details and quality control parameters of each European cohort included in Ishigaki *et al* (2022) meta-analysis have been previously described[3] and summarised in Supplementary Tables 1 and 2. Briefly, genotyping across the 25 European cohorts was performed using a range of Illumina arrays (including OmniExpress, HumanHap, HumanCNV, HumanCore/CoreExome, GSA, MEGA, and ImmunoChip platforms) and Affymetrix Genome-Wide SNP Arrays (versions 5.0 and 6.0). Samples with low call rate, closely related individuals and outliers in terms of ancestries were removed, as well as variants with low call rate, low MAF or low P-value for Hardy-Weinberg equilibrium (specific thresholds used in each cohort are detailed in Supplementary Tables 1 and 2)[3]. Genotypes were pre-phased using Shapeit2 or Eagle, and imputation was carried out using the 1000 Genomes Project Phase 3 reference panel with poorly imputed variants ($r^2 < 0.3$) removed[3]. Association tests were performed separately within each European cohort using PLINK (v2) with sex and genotype PCs as covariates (Supplementary Tables 1 and 2)[3]. Posterior, meta-analyses across all European cohorts were conducted using METAL[6] (inverse variance weighted fixed-effect model) for each trait.

### Supplementary Figures

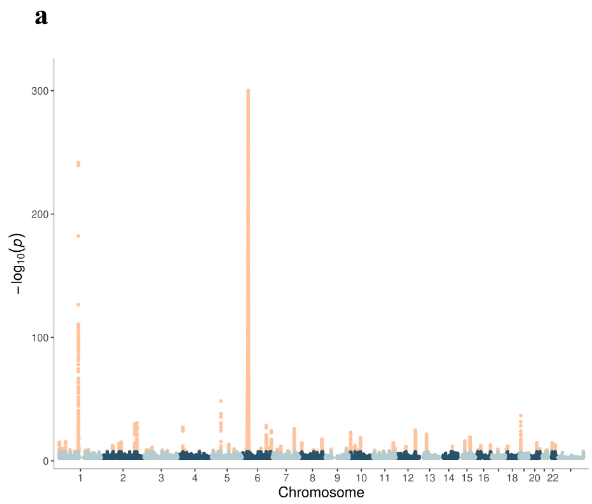


**Supplementary Figure 1 | Leave-one-cohort-out genome-wide association study meta-analysis results for rheumatoid arthritis phenotypes.** Manhattan plot of the rheumatoid arthritis (RA) (**a**) and seropositive RA (**b**) genome-wide association study meta-analyses, leaving Estonian Biobank out (*N* = 798,099 for RA and *N* = 795,183 for seropositive RA). Significant signals (over the Bonferroni corrected p-value of 5 x $10^{-8}$) are represented in orange, while non-significant ones are represented in blue and grey.

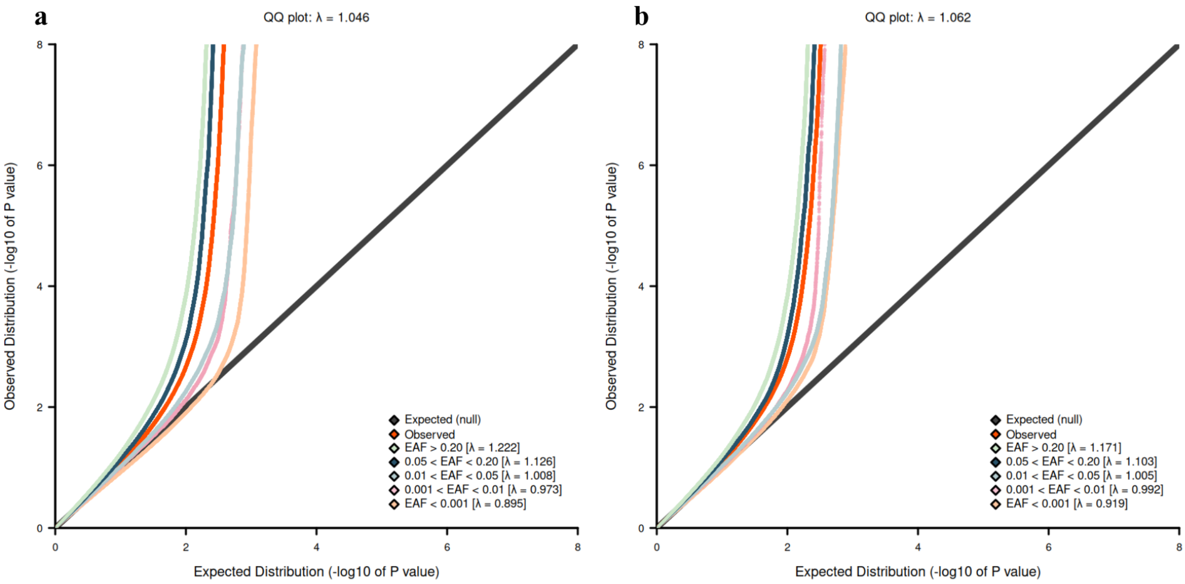


**Supplementary Figure 2 | Genome-wide association study meta-analysis results for rheumatoid arthritis phenotypes.** QQ plot of the rheumatoid arthritis (RA) (**a**) and seropositive RA (**b**) genome-wide association study meta-analyses (*N* = 1,000,776 for RA and *N* = 1,000,292 for seropositive RA). EAF, effect allele frequency.

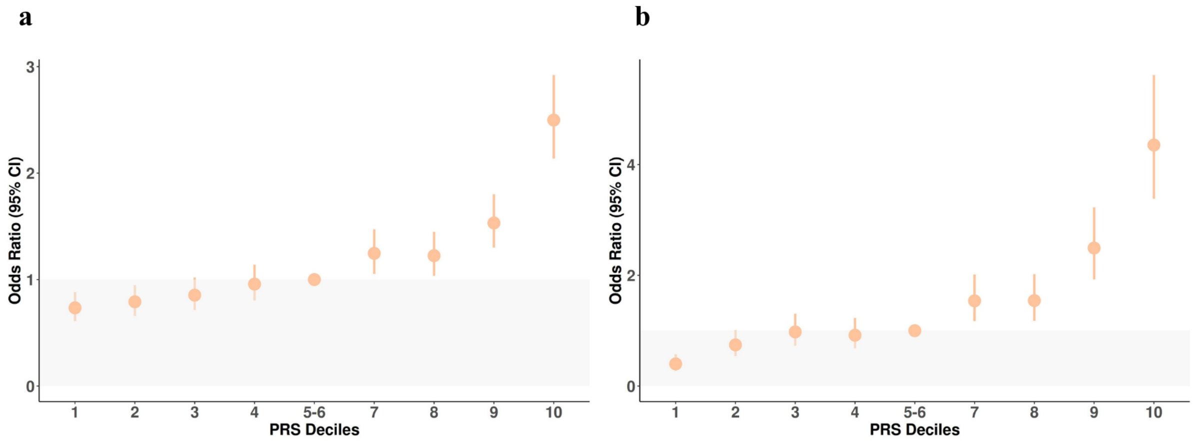


**Supplementary Figure 3 | Odds ratio across deciles.** Odds ratios (ORs) for prevalent rheumatoid arthritis (RA) (**a**) and seropositive RA (**b**) across polygenic risk score (PRS) deciles. Each decile represents 10% of the study population grouped by increasing genetic risk, with error bars indicating 95% confidence intervals (CIs). Grey area indicates OR < 1 relative to the reference deciles (5$^{th}$ and 6$^{th}$).

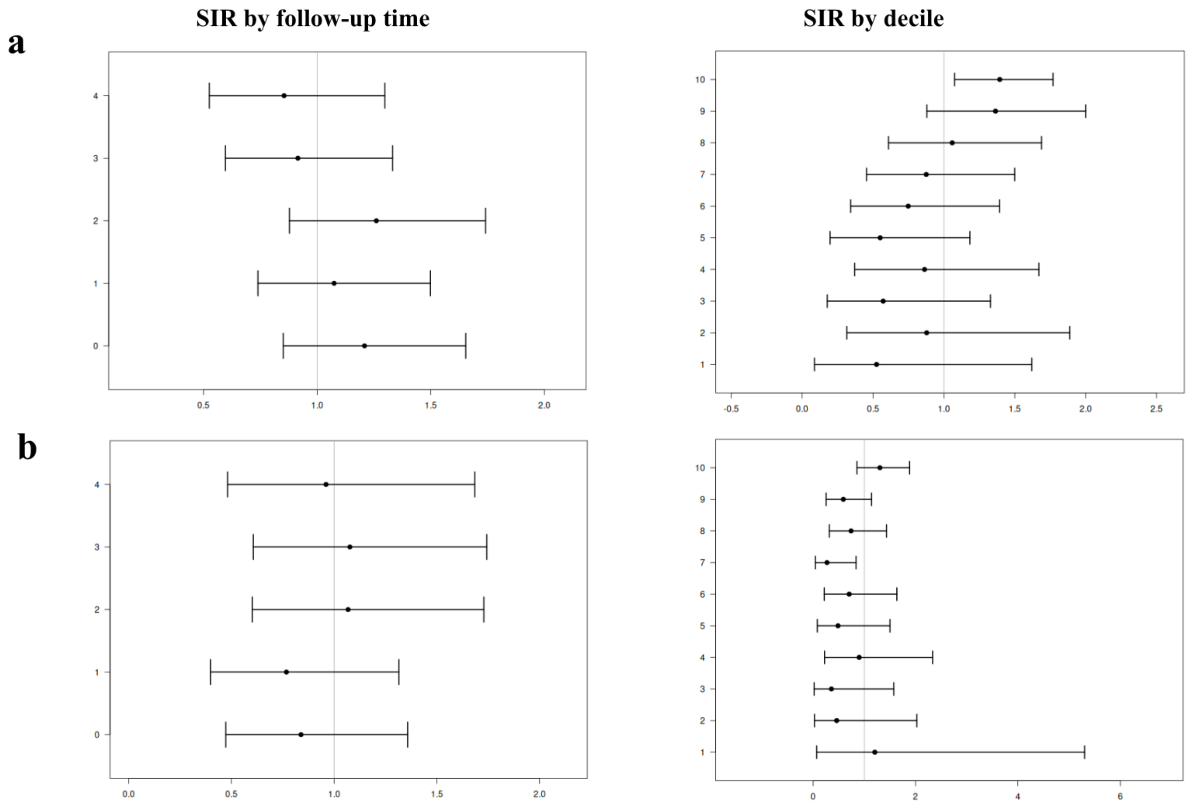


**Supplementary Figure 4 | Standardised incidence ratio of risk prediction models for rheumatoid arthritis phenotypes.** Standardised incidence ratio (SIR), adjusted by age, calendar year and sex, with 95% confidence intervals of risk prediction models for rheumatoid arthritis (RA) (**a**) and seropositive RA (**b**), assessed over five years of follow-up. SIRs are shown by follow-up time (**left**) and by risk decile (**right**).

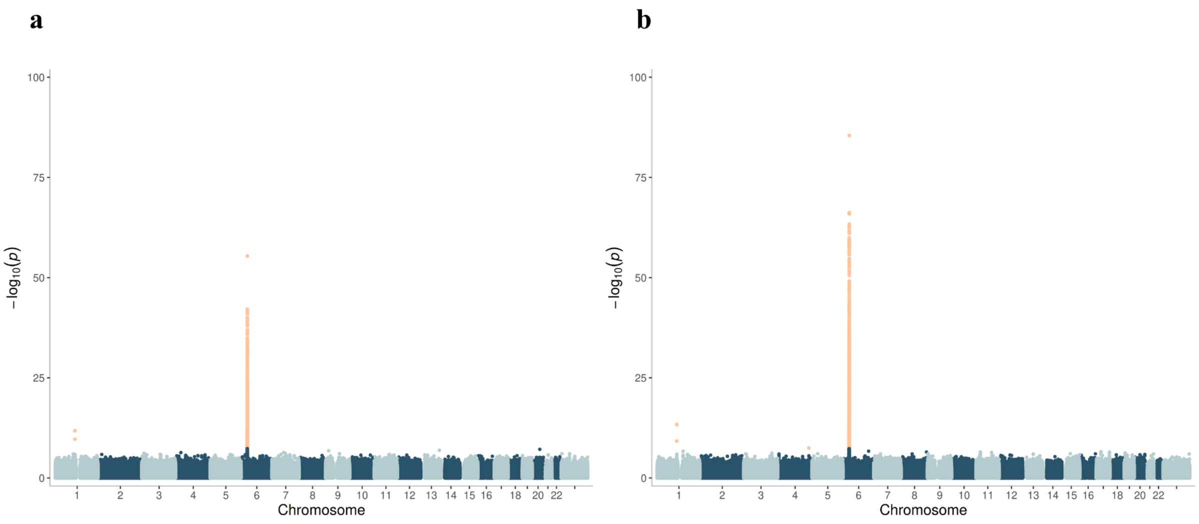


**Supplementary Figure 5 | Genome-wide association study results for rheumatoid arthritis phenotypes.** Manhattan plot of the rheumatoid arthritis (RA) (**a**) and seropositive RA (**b**) genome-wide association study ($N$ = 202,677 for RA and $N$ = 205,109 for seropositive RA) in Estonian Biobank. Significant signals (over the Bonferroni corrected p-value of 5 x $10^{-8}$) are represented in orange, while non-significant ones are represented in blue and grey.
